# Preliminary Evidence of a Link between COVID-19 Vaccines and Otologic Symptoms

**DOI:** 10.1101/2022.02.23.22271144

**Authors:** Ramsi A. Woodcock, Loren J. Bartels

**Author notes:** Cell Phone (preferred). Contribution: conceptualization; data curation; formal analysis; investigation; methodology; project administration; resources; software; supervision; validation; visualization; writing—original draft; writing—review & editing. *This author is not a medical doctor. His contributions to this paper are not intended to be, nor should they be taken as, medical advice*. Contribution: methodology; writing—review & editing (Loren J. Bartels).

## Abstract

**Hypothesis:** This study investigates whether U.S. Centers for Disease Control and Prevention Vaccine Adverse Events Reporting System (VAERS) data suggest an association between vertigo, tinnitus, hearing loss, Bell’s palsy and the COVID-19 vaccines administered in the United States.

**Background:** Published case reports suggest a possible association between various otologic symptoms and the COVID-19 vaccines, but the only published analysis of VAERS data, which did not account for underreporting of late-appearing adverse events, found no association between hearing loss and the vaccines.

**Methods:** The incidence in VAERS of vertigo, tinnitus, hearing loss, and Bell’s palsy associated with COVID-19 vaccinations administered between December 14, 2020 and June 7, 2021 was compared with published rates for the general population. To account for underreporting of late-appearing adverse events, incidences were calculated using only the initial part of the observation period, during which reported events spike above expected events.

**Results:** The COVID-19 vaccines were associated with statistically significant increases in the incidence of vertigo, tinnitus, hearing loss, and Bell’s palsy of 1877, 50, 12, and 14 cases per 100,000, respectively. In relation to the mRNA-1273 or BNT162b2 vaccines, the Ad26.COV2.S vaccine was associated with a statistically significant excess incidence of vertigo, tinnitus, and hearing loss of at least 723, 57, and 55 cases per 100,000, respectively.

**Conclusion:** These results suggest an association between the COVID-19 vaccines and vertigo, tinnitus, hearing loss, and Bell’s palsy. They also suggest that, with respect to vertigo, tinnitus, and hearing loss, the association is relatively strong for the Ad26.COV2.S vaccine.

## I. Introduction

Published case reports suggest a possible association between the COVID-19 vaccines and various otologic symptoms, including hearing loss, tinnitus, and vertigo.^1–3^ However, the only preliminary study of otologic symptoms in U.S. Centers for Disease Control and Prevention Vaccine Adverse Event Reporting System (VAERS) data published to date, in which Formeister et al. investigated reports of hearing loss, failed to find an association.^4,5^ This may be because vaccine adverse event reports relate to a single trigger—vaccination—whereas the population incidence rates to which they must be compared relate to multiple triggers.^6^ For example, the population incidence of vertigo includes cases of vertigo due to trauma, viral infection, and genetic disorders, among other things, whereas doctors and patients likely tend to report to VAERS only cases of vertigo that they believe to be triggered by vaccination. As a result, the incidence rate of vertigo in the VAERS reports will be lower than the population incidence rate unless vaccination triggers more vertigo than all other triggers combined.

The U.S. Centers for Disease Control and Prevention (CDC) tries to address this problem by encouraging doctors and patients to report all symptoms they encounter after vaccination.^7^ In theory, that should ensure that symptoms due to all triggers, including vaccination, are reported, with the result that, if vaccination triggers additional cases of a particular symptom, then the incidence rate in the VAERS reports will be higher than the population incidence rate and researchers will be able to infer from this difference that there is an association between the symptom and vaccination.

Unfortunately, doctors and patients do not heed the CDC. As Figure 1 shows, VAERS reports for all vaccines, COVID or otherwise, decline dramatically as the date of onset of the reported symptom extends beyond the vaccination date. If doctors and patients were to report all symptoms based on all triggers, including post-vaccination triggers, one would expect the line to be approximately flat, unless vaccination is a disproportionate source of all symptoms experienced by the population due to any cause, which is unlikely. In the absence of any test enabling doctors and patients to determine whether a particular symptom is caused by vaccination, doctors and patients rely on proximity of the date of symptom onset to the date of vaccination as a proxy for causation, which explains why, in Figure 1, reports spike for symptoms with onset immediately after vaccination and then fall off almost to zero after a few weeks.^8^ As shown in Figure 3, VAERS reports for the symptoms considered in this study—vertigo, tinnitus, hearing loss, and Bell’s palsy—all exhibit a similar initial spike in reported symptoms with onset immediately after vaccination, followed by a precipitous decline in reported symptoms as the date of onset moves farther away from the date of vaccination.

**Figure 1:**
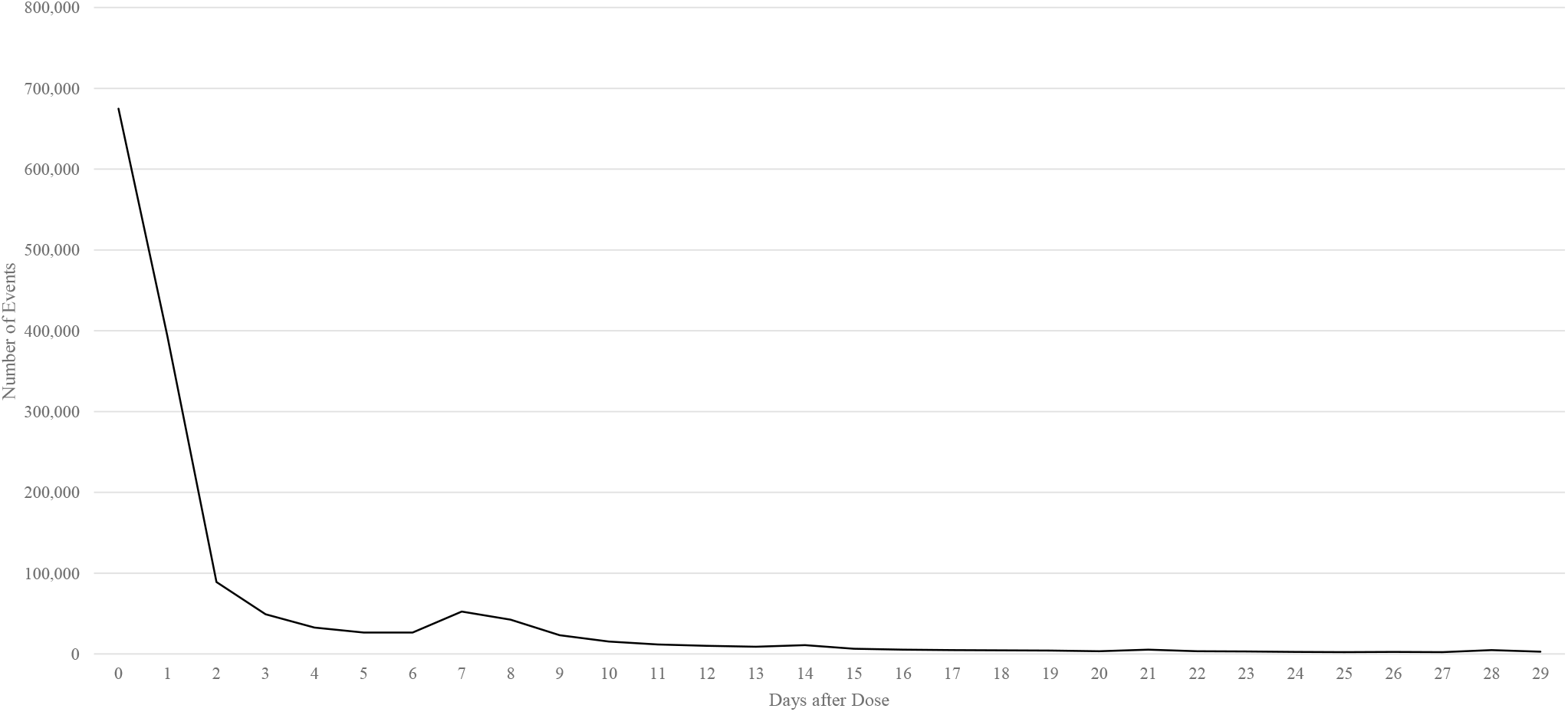
The Early-Onset Spike in VAERS Reports: All Adverse Events Reported to VAERS for All Vaccines, COVID-19 and Non-COVID-19, Administered between December 14, 2020 and June 7, 2021.

One way around the failure of doctors and patients fully to report symptoms due to non-vaccine triggers would be to use published reporting sensitivity rates to inflate the raw number of VAERS reports. Unfortunately, existing studies of VAERS reporting sensitivity, such as that of Miller et al., are based on VAERS reports that have been selected for their temporal proximity to vaccination—the same filter that doctors and patients place on their own reporting—and hence are unlikely fully to capture underreporting of symptoms associated with non-vaccine triggers.^9^ Sensitivity rates extrapolated from a comparison with clinical trial adverse event reports suffer from a similar limitation, because clinical trials solicit reports only within a few days of vaccination.

Fortunately, there is another way around the underreporting of non-vaccine-triggered events. If the number of cases reported during the peak reporting period immediately after vaccination— the “spike period”—exceeds the number of cases expected during that period based on the population incidence rate of the symptom, then it follows that the vaccine is a trigger of the symptom, even if reported cases fall below expected cases for the rest of the observation period, and even if the incidence rate implied by the reports, if calculated over the entire observation period, is below the population incidence rate. For the only way in which reported cases can exceed expected cases during any part of an observation period, despite underreporting of cases due to non-vaccine triggers, is if vaccine-triggered cases of the symptom are numerous enough over that period to make up the difference. But the existence of any number of vaccine-triggered cases, if statistically significant in magnitude, is sufficient to establish an association between the vaccine and the symptom.

Consider a numerical illustration. Suppose that vaccination triggers one hundred excess cases of vertigo in the first day after vaccination and none on the second day, and that all other triggers cause an additional hundred cases every day, but that doctors and patients, despite reporting all symptoms on the first day regardless of trigger, report none on the second day, because they assume that none of the day-two symptoms are triggered by the vaccine. If 365,000,000 vaccinations are observed, then for a 2-day observation period, the incidence of vertigo is 10 (per 100,000 person-years, as with all incidence rates discussed in the present study), but the population incidence (which is based on the number of cases caused by all other triggers) is 15, suggesting, falsely, that there is no association between vaccination and the symptom. If, however, one considers only the first day, during which total reported cases of 200 spike above expected cases of 100, then the incidence is 20, which exceeds the population incidence of 15 and therefore correctly indicates the existence of an association.

An additional consideration complicates analysis of spike periods: reports may not be accurate. As a result, the presence of a spike in raw report numbers for a particular symptom is not sufficient to suggest the existence of an association between the vaccine and the symptom. The spike must be discounted, either through examination of individual reports or by a rate of accuracy derived from analysis of past reports. Even if discounting eliminates the spike, however, the analysis does not end, because the studies, such as that of Miller et al., that provide sensitivity rates for reporting of adverse events with onset close to the date of vaccination are likely to capture underreporting during spike periods even if they do not capture underreporting of cases thereafter.^9^ The same is true for sensitivity rates extrapolated from a comparison with clinical trial reports. Published sensitivity rates and those extrapolated from clinical trials can therefore be used to offset the accuracy discount with a multiplier that accounts for underreporting during the spike.

## II. Materials & Methods

To investigate the relationship between the COVID vaccines administered in the United States (Moderna’s mRNA-1273, Pfizer-BioNTech’s BNT162b2, and Johnson & Johnson’s Ad26.COV2.S) and otologic symptoms, analysis of spikes in reports to VAERS of vertigo, tinnitus, hearing loss, and Bell’s palsy over expected levels were carried out. To avoid bias associated with the ongoing arrival of new reports regarding the COVID-19 vaccines, the analysis was limited to vaccines administered between December 14, 2020 and June 7, 2021, to symptoms with an onset within 30 days of vaccination, and to reports made within 60 days of vaccination. The MedDRA-coded symptom fields in the VAERS database were searched for “vertigo,” “tinnitus,” “hearing loss,” “bell’s palsy,” and synonyms, and the results filtered for the “COVID19” vaccine type to create datasets for each symptom. Table 1 and Figure 2 provide more information on the search terms, datasets, and study parameters.

**Table 1:** Overview of the Data Used in the Study.

| Table 1: Overview of the Data Used in the Study. |  |  |
| --- | --- | --- |
| Study parameters | Period over which vaccinations included in the study were administered | December 14, 2020 to June 7, 2021 |
|  | Days after vaccination observed | 30 |
|  | Days after vaccination during which adverse event reports were collected | 60 |
| Vertigo<sup>1</sup> cases reported to VAERS (search term: “vertigo”) | **Total cases** | **3,993** |
|  | mRNA-1273 | 1,713 |
|  | BNT162b2 | 1,950 |
|  | Ad26.COV2.S | 323 |
| Tinnitus<sup>2</sup> cases reported to VAERS (search terms: “tinnitus”; “hypoacusis”) | **Total cases** | **6,562** |
|  | mRNA-1273 | 2,615 |
|  | BNT162b2 | 3,366 |
|  | Ad26.COV2.S | 573 |
| Hearing loss<sup>3</sup> cases reported to VAERS (search terms: “hearing loss”; “deafness”; “hypoacusis”) | **Total cases** | **1,962** |
|  | mRNA-1273 | 750 |
|  | BNT162b2 | 1,036 |
|  | Ad26.COV2.S | 170 |
| Bell’s palsy<sup>4</sup> cases reported to VAERS (search terms: “bell’s palsy”; “facial palsy”; “facial paralysis”) | **Total cases** | **3,541** |
|  | mRNA-1273 | 1,463 |
|  | BNT162b2 | 1,824 |
|  | Ad26.COV2.S | 248 |
| Overall vaccinated population (at least one dose)<sup>5</sup> | **Total** | **171,310,738** |
|  | mRNA-1273 | 69,355,687 |
|  | BNT162b2 | 90,933,285 |
|  | Ad26.COV2.S | 11,021,766 |

<sup>1</sup> Cases by manufacturer do not sum to total cases because the vaccine manufacturer was not identified in seven cases.
<sup>2</sup> Cases by manufacturer do not sum to total cases because the vaccine manufacturer was not identified in eight cases.
<sup>3</sup> Cases by manufacturer do not sum to total cases because the vaccine manufacturer was not identified in six cases.
<sup>4</sup> Cases by manufacturer do not sum to total cases because the vaccine manufacturer was not identified in six cases.
<sup>5</sup> The manufacturer figures have been scaled to equal the total number of first-doses reported by the CDC to have been administered. It is not clear why the manufacturer figures reported by the CDC do not sum to the total without scaling.

**Figure 2:**
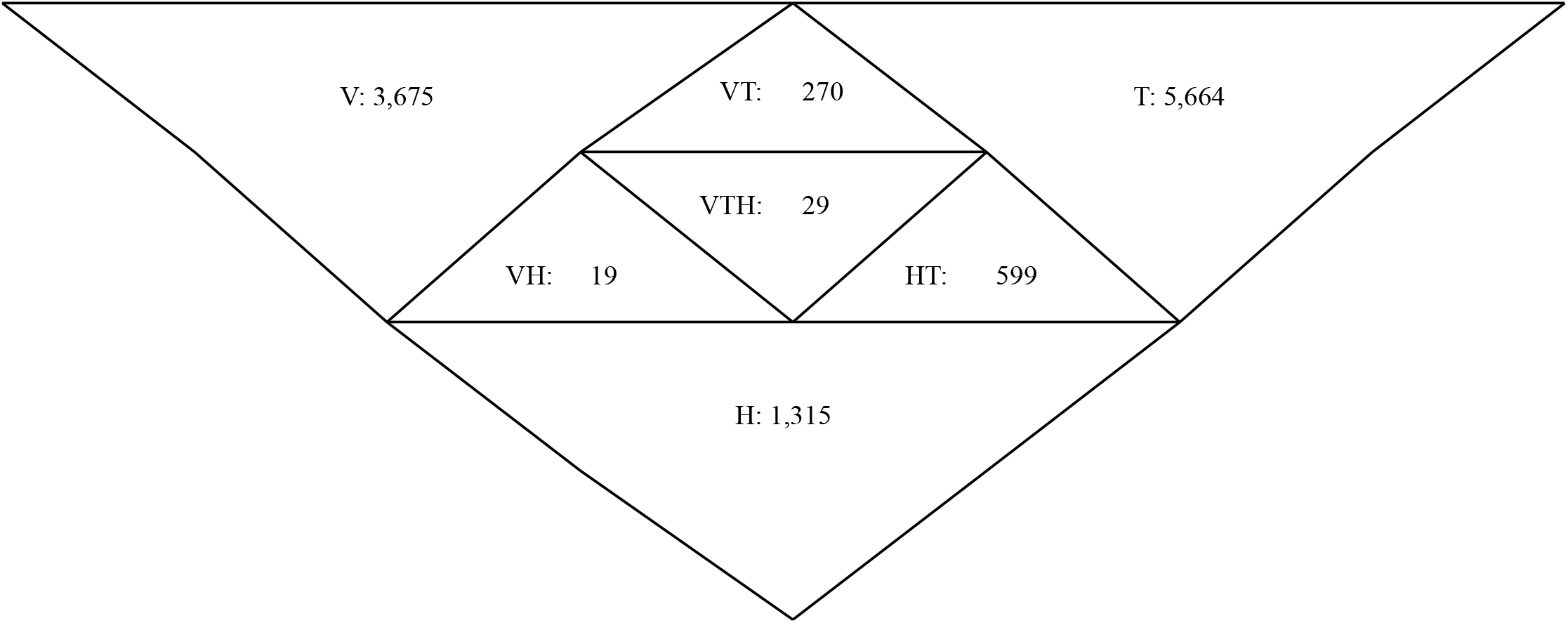
Triangle Venn Diagram Showing the Intersection of the Sets of Reports of Vertigo (V), Tinnitus (T), and Hearing Loss (H) in VAERS data.

Formeister et al. manually reviewed each VAERS adverse event report in their study sample for accuracy. For the present study of multiple symptoms relating to nearly seven months of vaccinations, as opposed to the study of a single symptom over 2.5 months carried out by Formeister et al., manual verification could not be undertaken. Instead, a best-guess accuracy rate was identified by running the same search of the VAERS data for hearing loss reported by Formeister et al. and using the resulting case count of 475 to extrapolate an accuracy rate of 8% from Formeister et al.’s conclusion that only 40 reported cases in their VAERS data were accurate. This appears to be a very low accuracy rate. By contrast, an accuracy rate of 57% is implied by the work of Su et al. on anaphylaxis reports to VAERS in relation to a number of non-COVID-19 vaccines.^10^

A reporting sensitivity rate of 30%, corresponding to a multiplier of 3.3, was used for tinnitus, hearing loss, and Bell’s palsy. This rate was obtained by averaging the rates reported by Miller et al. for reports of anaphylaxis and Guillain-Barré syndrome associated with non-COVID-19 vaccinations.^9^ It is somewhat lower than the 50% sensitivity rate that appears to have been arbitrarily chosen for sensitivity testing of hearing loss reports by Formeister et al.

A much lower sensitivity rate—0.6%, corresponding to a multiplier of 166.9—was used for vertigo, because, as Neuhauser has pointed out, vertigo is common in the general population, affecting between 20% and 30% of adults.^11^ Doctors and patients may therefore be less likely to report vertigo in connection with the COVID-19 vaccines than they are to report tinnitus, hearing loss, or Bell’s palsy, each of which is rare in comparison.^11^ To identify an appropriate reporting sensitivity rate for vertigo, the incidence of other common symptoms—pain, erythema, swelling, fever, headache, fatigue, myalgia, and nausea or vomiting—reported in the COVID-19 vaccines’ Phase III clinical trials was compared with the incidence of the same symptoms in the VAERS reports and the largest result—that for fever—was used to obtain the 0.6% reporting sensitivity rate.^12–14^ A comparison with clinical trial reports was considered probative of the true reporting sensitivity rate during the spike period because clinical trial administrators solicit reports from trial subjects within seven days of vaccination.^12–14^

Spike periods were determined after application of the 8% accuracy discount and the applicable underreporting multiplier to the VAERS report counts for each symptom. As shown in Figure 3 and discussed more fully in the Results section, the spike period was two days each for vertigo, tinnitus, and Bell’s palsy, and one day for hearing loss. The statistical significance of the incidence rate of each symptom over its spike period, in relation to the published population incidence rate for the symptom, was determined using the normal approximation to the binomial distribution for one-sample inference with two tails.^15–18^

**Figure 3:**
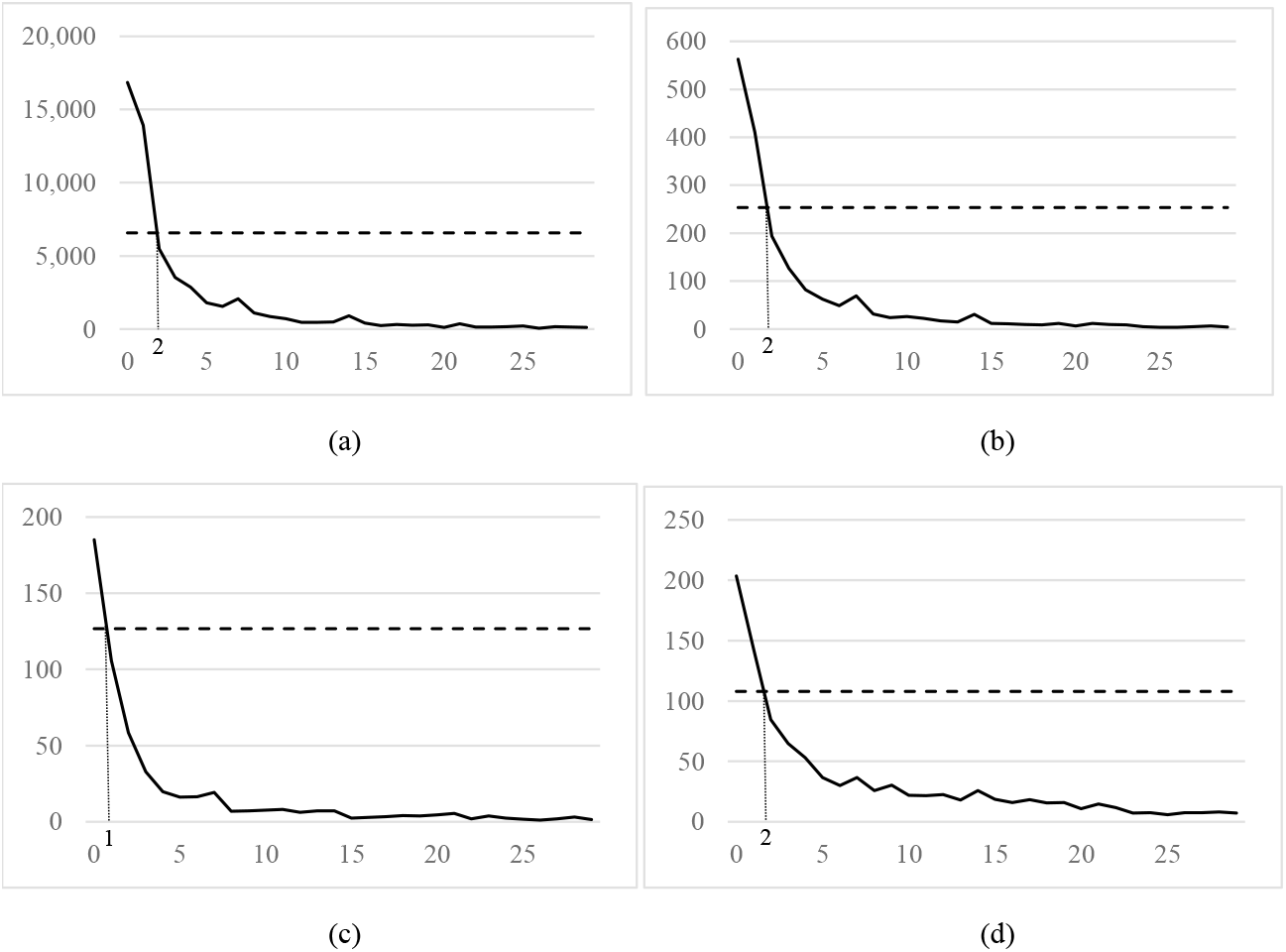
Spike Periods for Otologic Symptoms: Reports to VAERS of (a) Vertigo, (b) Tinnitus, (c) Hearing Loss, and (d) Bell’s Palsy in Relation to the COVID-19 Vaccines for Doses Administered between December 14, 2020 and June 7, 2021, Showing Spike Periods During Which Reported Cases Exceed Expected Cases. The solid line shows the number of cases reported and the horizontal dashed line shows the expected number of cases based on the population incidence rate. The vertical dotted line shows the number of days after dose administration at which the spike of reported cases above expected cases ends. The vertical axis is labeled in report numbers and the horizontal access is labeled in the number of days between administration of the dose and symptom onset. An accuracy discount rate of 8% has been applied to all case counts, an underreporting multiplier of 3.3 has been applied to the tinnitus, hearing loss, and Bell’s palsy case counts, and an underreporting multiplier of 166.9 has been applied to the vertigo case counts.

The proportional reporting ratio (PRR),^19^ which compares the incidence rate of symptoms in reports related to the vaccine at issue (here, the COVID-19 vaccines) to the rate of all other symptoms in all other VAERS reports,^20^ was also calculated for all four symptoms. The PRR has two helpful features. First, it compares VAERS data with other VAERS data, so the problem of comparing single-trigger VAERS incidence rates to multiple-trigger population incidence rates is avoided. Second, the PRR provides a useful check on the appropriateness of the accuracy and reporting sensitivity rates used in the present study, because, if accuracy and reporting sensitivity rates are constant across all vaccines, then these rates cancel out of the PRR, making the PRR invariant in reporting accuracy and sensitivity assumptions. The statistical significance of the PRRs, in relation to the null hypothesis of one (i.e. that there is no difference between the incidence of the symptom in COVID-19 vaccine reports and the incidence of all other symptoms in all other vaccine reports) was determined using the chi-squared test with one degree of freedom.

Finally, to investigate the relative association of different COVID-19 vaccines with vertigo, tinnitus, hearing loss, and Bell’s palsy, VAERS spike-period case counts, discounted as above, were categorized by vaccine manufacturer, incidence rates for each were calculated using CDC data on total administered vaccine doses per manufacturer, and, for each symptom, the normal approximation to the binomial distribution for two-sample inference was applied with two tails. For all three tests, p-values below 0.05 were considered significant.

University of Kentucky determined that this study is exempt from institutional review board approval because the study uses deidentified data publicly available on the CDC website.

## III. Results

Figure 3 shows the distribution of cases, after adjustment for accuracy and underreporting, in terms of time to symptom onset, including the spike period for each symptom. The incidence rate of vertigo in the VAERS reports over vertigo’s two-day spike period was 3,277 (per 100,000 person-years, as for all incidence rates reported here), significantly in excess of the population incidence of vertigo of 1,400 reported by Neuhauser et al. (p < 0.00001).^17^ The incidence rate of tinnitus in the VAERS reports over tinnitus’s two-day spike period was 104, significantly in excess of the population incidence of tinnitus of 54 reported by Martinez et al. (p < 0.00001).^16^ The incidence rate of hearing loss in the VAERS reports over hearing loss’s one-day spike period was 39, significantly in excess of the population incidence of hearing loss of 27 reported by Alexander et al. (p < 0.00001).^18^ The incidence rate of Bell’s palsy in the VAERS reports over Bell’s palsy’s two-day spike period was 37, significantly in excess of 23, which is the average of the population incidence rates for Bell’s palsy referenced by Ozonoff et al. (p < 0.00001).^15^ As Table 2, which reports these results, shows, incidence rates calculated using the three-week observation period employed by Formeister et al., rather than the spike periods, were, by contrast, all lower than their respective population incidence rates, by 846 for vertigo, 36 for tinnitus, 22 for hearing loss, and 14 for Bell’s palsy. Incidence rates in the VAERS reports break even with population incidence rates at approximately a one-week observation period.

**Table 2:** Comparing Spike Period Incidence Rates to General Population Incidence Rates. All incidence rates are annual per 100,000 persons.

Tables for “Preliminary Evidence of a Link between COVID-19 Vaccines and Otologic Symptoms”
|  |  |  |  |
| --- | --- | --- | --- |
| Symptom | Cases During Spike Period /<br>Expected Cases for Spike<br>Period / Spike Period (Days) | Incidence over Spike Period (95% CI)<br>/ General Population Incidence /<br>Incidence over 21-Day Observation<br>Period | z-statistic / p-value (two-tailed) / significant at the<br>95% confidence level? |
| Accuracy: 8%; Reporting Sensitivity/Multiplier: 30%/3.3 (vertigo: 0.6%/166.9); Overall: 28% (vertigo: 1,405%) |  |  |  |
| Vertigo | 30,760.64 / 13,141.65 / 2 | 3,276.98 (3,240.96, 3,313.00) / 1,400 /<br>554.47 | 154.78 / <0.00001 / **yes** |
| Tinnitus | 972.94 / 506.89 / 2 | 103.65 (97.14, 110.16) / 54 / 18.06 | 20.71 / <0.00001 / **yes** |
| Hearing<br>Loss | 185.07 / 126.72 / 1 | 39.43 (33.75, 45.11) / 27 / 5.34 | 5.18 / <0.00001 / **yes** |
| Bell's<br>Palsy | 346.06 / 215.90 / 2 | 36.87 (32.98, 40.75) / 23 / 9.27 | 8.86 / <0.00001 / **yes** |
| Accuracy: 8%; Reporting Sensitivity/Multiplier: 50%/2 (vertigo: 1%/100); Overall: 17% (vertigo: 845%) |  |  |  |
| Vertigo | 18,503.71 / 13,141.65 / 2 | 1,971.23 (1,943.11, 1,999.35) / 1,400 /<br>333.53 | 47.11 / <0.00001 / **yes** |
| Tinnitus | 338.53 / 253.45 / 1 | 72.13 (64.45, 79.81) / 54 / 10.87 | 5.35 / <0.00001 / **yes** |
| Bell's<br>Palsy | 122.44 / 107.95 / 1 | 26.09 (21.47, 30.71) / 23 / 5.58 | 1.40 / 0.16 / no |
| Hearing<br>Loss | No spike period; reported cases never exceed expected cases, suggesting the absence of a positive association between hearing loss and the COVID-19 vaccines. Incidence over a 21-day observation period is 3.21 per 100,000 person-years. |  |  |

The results for vertigo and tinnitus were robust to lower underreporting multipliers. As Table 2 also reports, a statistically significant excess incidence of tinnitus remained after reduction of the underreporting multiplier from 3.3 to the value of 2 used by Formeister et al., and a statistically significant excess incidence of vertigo remained after the underreporting multiplier was scaled down in the same proportion (i.e., by the ratio of 2 to 3.3).

The PRRs, which require no assumption regarding the rate of accuracy or the rate of reporting sensitivity, were, for vertigo, tinnitus, hearing loss, and Bell’s palsy, 4.76, 5.41, 2.63, and 1.46, respectively, as shown in Table 3. Those for vertigo and tinnitus exceeded the null hypothesis of one to a statistically significant degree (p < 0.001 for vertigo and p < 0.0001 for tinnitus). This result was consistent with the robustness of the spike period results for vertigo and tinnitus.

**Table 3:** Proportional Reporting Ratios for Vertigo, Tinnitus, Hearing Loss, and Bell’s Palsy.

Tables for “Preliminary Evidence of a Link between COVID-19 Vaccines and Otologic Symptoms”
|  |  |  |  |  |  |
| --- | --- | --- | --- | --- | --- |
| Variable | (a) Cases after a COVID-19 vaccine dose | (b) Reports of all other adverse events after a COVID-19 vaccine dose | (c) Cases after all other non-COVID-19 vaccine doses | (d) Reports of all other adverse events after all other non-COVID-19 vaccine doses | PRR $\frac{\frac{a}{a+b}}{\frac{c}{c+d}}$ / $\chi^2$ statistic (1 df) / p-value / significant at the 95% confidence level? |
| Vertigo | 3,993 | 458,837 | 9 | 4,955 | 4.76 / 14.13 / <0.001 / **yes** |
| Tinnitus | 6,562 | 456,268 | 13 | 4,951 | 5.41 / 19.48 / <0.0001 / **yes** |
| Hearing Loss | 1,962 | 460,868 | 8 | 4,956 | 2.63 / 2.66 / 0.103 / **no** |
| Bell’s Palsy | 3,541 | 459,289 | 26 | 4,938 | 1.46 / 0.21 / 0.65 / **no** |

These results were clinically significant. As Table 2 shows, over the course of the two-day spike period after vaccination, 30,761 of the 171 million people who received at least one dose of vaccine in the United States during the study period (about 1 per 5,569 persons receiving at least one dose) are estimated to have had vertigo. That was 17,619 more cases than the 13,142 cases that were expected over that period based on the population incidence of vertigo (an excess rate of 1 per 9,723 persons receiving at least one dose). For tinnitus, the excess of reported cases over expected cases over the two-day tinnitus spike period was 466 (an excess rate of 1 per 367,579 persons receiving at least one dose). For hearing loss, it was 58 excess cases over the one-day hearing loss spike period (about 1 per 2.94 million persons receiving at least one dose), and for Bell’s palsy it was 130 excess cases over the two-day Bell’s palsy spike period (about 1 extra Bell’s palsy case per 1.32 million persons receiving at least one dose).

Table 4 sets forth the results regarding the relative associations of the three vaccines administered in the United States. The incidence of vertigo associated with the Ad26.COV2.S vaccine in the VAERS reports was 4,258, in excess of the incidence of 3,535 associated with the mRNA-1273 vaccine and the incidence of 2,947 associated with the BNT162b2 vaccine. The excess in the incidence of vertigo associated with the Ad26.COV2.S vaccine in relation to both the mRNA-1273 vaccine and the BNT162b2 vaccine was statistically significant (p < 0.00001), as was the excess in the incidence of vertigo associated with the mRNA-1273 vaccine in relation to the BNT162b2 vaccine (p < 0.00001). The incidence of tinnitus associated with the Ad26.COV2.S vaccine in the VAERS reports was 157, in excess of the incidence of 100 associated with both the mRNA-1273 and BNT162b2 vaccines to a statistically significant degree (p < 0.001 and p<0.0001, respectively). The incidence of hearing loss associated with the Ad26.COV2.S vaccine in the VAERS reports was 93, in excess of the incidences associated with the mRNA-1273 and BNT162b2 vaccines of 33 and 37, respectively, to a statistically significant degree (p < 0.00001 and p < 0.0001, respectively). The incidence of Bell’s palsy associated with the BNT162b2 and mRNA-1273 vaccines in the VAERS reports was 37 for both, which exceeded the incidence of 36 associated with the Ad26.COV2.S vaccine, but not to a statistically significant degree.

**Table 4:** Relative Incidence of Vertigo, Tinnitus, Hearing Loss, and Bell’s Palsy Associated with the mRNA-1273, BNT162b2, and Ad26.COV2.S COVID-19 Vaccines. All incidence rates are annual per 100,000 persons.

Tables for “Preliminary Evidence of a Link between COVID-19 Vaccines and Otologic Symptoms”
|  |  |  |  |  |  |
| --- | --- | --- | --- | --- | --- |
| Accuracy: 8%; Reporting Sensitivity/Multiplier: 30%/3.3 (vertigo: 0.6%/166.9); Overall: 28% (vertigo: 1,405%) |  |  |  |  |  |
| Adverse event | Vaccine | Cases During Spike Period / Total Recipients over Full Observation Period | Incidence over Spike Period / General Population Incidence | Comparison with mRNA-1273 Vaccine: z-statistic / p-value (two-tailed) / Significant at the 95% confidence level? | Comparison with BNT162b2 Vaccine: z-statistic / p-value (two-tailed) / Significant at the 95% confidence level? |
| Vertigo | mRNA-1273 | 13,434.07 / 69,355,687 | 3,534.99 / 1,400 | n/a | 15.50 / <0.00001 / **yes** |
|  | BNT162b2 | 14,684.73 / 90,933,285 | 2,947.17 / 1,400 | n/i | n/a |
|  | Ad26.COV2.S | 2,571.58 / 11,021,766 | 4,258.07 / 1,400 | 8.81 / <0.00001 / **yes** | 17.57 / <0.00001 / **yes** |
| Tinnitus | mRNA-1273 | 379.94 / 69,355,687 | 99.98 / 54 | n/a | 0.0 / 1.0 / no |
|  | BNT162b2 | 497.53 / 90,933,285 | 99.85 / 54 | n/i | n/a |
|  | Ad26.COV2.S | 94.63 / 11,021,766 | 156.70 / 54 | 3.88 / <0.001 / **yes** | 3.99 / <0.0001 / **yes** |
| Hearing Loss | mRNA-1273 | 63.28 / 69,355,687 | 33.30 / 27 | n/a | n/i |
|  | BNT162b2 | 93.23 / 90,933,285 | 37.42 / 27 | 0.64 / 0.52 / no | n/a |
|  | Ad26.COV2.S | 28.00 / 11,021,766 | 92.72 / 27 | 4.56 / <0.00001 / **yes** | 4.21 / <0.0001 / **yes** |
| “ | “ | “ | “ | “ | Comparison with Ad26.COV2.S Vaccine: . . . |
| Bell’ s Palsy | mRNA-1273 | 139.15 / 69,355,687 | 36.62 / 23 | n/a | 0.0 / 1.0 / no |
|  | BNT162b2 | 184.51 / 90,933,285 | 37.03 / 23 | 0.04 / 0.96 / no | 0.05 / 0.96 / no |
|  | Ad26.COV2.S | 21.56 / 11,021,766 | 35.70 / 23 | n/i | n/a |

These results were also clinically significant. The comparatively higher vertigo incidence rate of the Ad26.COV2.S vaccine implies that, had the 171 million people who received at least one vaccine dose over the study period all received the Ad26.COV2.S vaccine, rather than one of the assortment of Ad26.COV2.S, BNT162b2, and mRNA-1273 vaccines that was actually administered, there would have been 39,970 cases of vertigo over the two-day spike period, 9,209 more than the 30,761 cases estimated to have actually occurred (an excess rate of about 1 per 18,602 persons receiving at least one dose). Similarly, the vertigo incidence rate of the mRNA-1273 vaccine, which was elevated relative to the vertigo incidence rate of the BNT162b2 vaccine, implies that, had mRNA-1273 alone been administered, there would have been 33,183 cases of vertigo over the two-day spike period, 2,422 more than the 30,761 cases estimated to have actually occurred (an excess rate of about 1 per 70,733 persons receiving at least one dose). The comparatively higher tinnitus incidence rate of the Ad26.COV2.S vaccine implies that, had Ad26.COV2.S alone been administered, there would have been 1,471 cases of tinnitus over the two-day spike period, 498 more than the 973 cases estimated to have actually occurred (an excess rate of 1 per 344,028 persons receiving at least one dose). Finally, the comparatively higher hearing loss incidence rate of the Ad26.COV2.S vaccine implies that, had Ad26.COV2.S alone been administered, there would have been 435 cases of hearing loss over the one-day spike period, 250 more than the 185 cases estimated to have actually occurred (an excess rate of 1 per 684,947 persons receiving at least one dose).

## IV. Discussion

These results suggest a statistically significant association between the COVID-19 vaccines and otologic symptoms, particularly vertigo and tinnitus, during the one-to-two-day immediate post-vaccination periods over which reported cases for these symptoms spike above expected cases. The results contrast with results obtained by including cases with later onset in the observation period, such as those published by Formeister et al. The results also suggest that the Ad26.COV2.S vaccine is more likely to trigger vertigo, tinnitus, or hearing loss than are the mRNA-1273 or BNT162b2 vaccines. The number of excess cases of vertigo, tinnitus, hearing loss, and Bell’s palsy associated with the vaccines is quite small relative to the total number of people who have received a dose of the vaccines, particularly in the case of tinnitus, hearing loss, and Bell’s palsy, but still clinically significant. The existence of an association over the one-to-two-day spike periods does not exclude the existence of an association between the vaccines and symptoms with later onset. Underreporting to VAERS prevents verification of such an association, however.

More investigation is required to confirm these associations, because the true accuracy and underreporting sensitivity rates are unknown. One reason for which the 30% sensitivity rate applied for tinnitus, hearing loss, and Bell’s palsy might be incorrect is that media attention paid to the COVID-19 vaccines may have pushed their reporting sensitivity rates above those of the vaccines considered by Miller et al., upon which the 30% rate is based.^9,21^ More investigation is also required to determine the extent of recovery from the symptoms considered in the present study, either to tolerability or to complete resolution, especially as there are reports of improvement for some symptoms.^2,3^

## V. Conclusion

This study created a novel way to compare the incidence of post-vaccine symptoms to published population incidence rates by using only cases with onset in the immediate one-to-two-day post-vaccination periods during which reported cases spike above expected cases to calculate incidence rates. In looking at VAERS reports of vertigo, tinnitus, hearing loss, and Bell’s palsy in recipients of COVID-19 vaccines, statistical evidence of an association of the vaccines with an excess of these cases appears at rates ranging from about 1 excess case per 5,500 vaccinated persons to about 1 excess case per 2.94 million vaccinated persons. Variations of this methodology might be helpful in investigating the risks of other vaccines.

## Data Availability

No data were produced for the study. All data analyzed in this study were downloaded from https://wonder.cdc.gov/vaers.html. All data and calculations used in the study can be downloaded in Excel format from this link: https://www.dropbox.com/s/82m4h7cba9g6h4v/TinnitusHearingLossVAERS%205.xlsx?dl=0

## Notes

Sources of Support/Disclosure of Funding: This study received no outside support or funding.

Conflicts of Interest: Ramsi A. Woodcock suffers from otologic symptoms that he believes to have been caused by COVID-19 vaccination. He is Loren J. Bartels’ patient.

### Competing Interest Statement

Ramsi A. Woodcock suffers from otologic symptoms that he believes to have been caused by COVID-19 vaccination. He is Loren J. Bartels' patient.

### Funding Statement

This study did not receive any funding.

### Author Declarations

The study uses data from the Vaccine Adverse Event Reporting System (VAERS) that is publicly available on the CDC website at https://wonder.cdc.gov/vaers.html.

